# Qualitative evaluation of the use of modelling in resource allocation decisions for HIV and TB

**DOI:** 10.1101/2023.04.11.23288405

**Authors:** Anna L Bowring, Debra ten Brink, Rowan Martin-Hughes, Nicole Fraser-Hurt, Nejma Cheikh, Nick Scott

## Abstract

**Introduction:** Globally, resources for health spending, including HIV and tuberculosis (TB), are constrained, and a substantial gap exists between spending and estimated needs. Optima is an allocative efficiency modeling tool that has been used since 2010 in over 50 settings to generate evidence for country-level HIV and TB resource allocation decisions. This evaluation assessed the utilization of modeling to inform financing priorities from the perspective of country stakeholders and their international partners.

**Methods:** In October-December 2021, the World Bank and Burnet Institute led 16 semi-structured small-group virtual interviews with 54 representatives from national governments and international health and funding organizations. Interviews probed participants’ roles and satisfaction with Optima analyses and how model findings have had been used and impacted resource allocation. Interviewed stakeholders represented nine countries and 11 different disease program-country contexts with prior Optima modeling analyses. Interview notes were thematically analyzed to assess factors influencing the utilization of modeling evidence in health policy and outcomes.

**Results:** Common influences on utilization of Optima findings encompassed the perceived validity of findings, health system financing mechanisms, the extent of stakeholder participation in the modeling process, engagement of funding organization, socio-political context, and timeliness of the analysis. Utilizing workshops can facilitate effective stakeholder engagement and collaboration.

Model findings were often used conceptually to localize global evidence and facilitate discussion. Secondary outputs included informing strategic and financial planning, funding advocacy, grant proposals, and influencing investment shifts.

**Conclusion:** Allocative efficiency modeling has supported evidence-informed decision making in numerous contexts and enhanced the conceptual and practical understanding of allocative efficiency. Most immediately, greater involvement of country stakeholders in modeling studies and timing studies to key strategic and financial planning decisions may increase the impact on decision making. Better consideration for integrated disease modeling, equity goals, and financing constraints may improve relevance and utilization of modeling findings.

---

**What is already known on this topic -**

Mathematical modeling is widely used in health planning and policy, including to support understanding of infectious disease epidemics at national and global levels. Allocative efficiency modeling tools such as Optima are used to consider the most cost-effective distribution of resources to maximize specified health gains. Many global institutions place high importance on evidence-informed policies, and there is an array of literature and conceptual frameworks describing the utilization of research evidence in policy and resulting outcomes. However, even though resource allocation is critical to HIV and TB program outcomes, there are limited examples of groups evaluating the utilization of modeling evidence to inform resource allocation.

**What this study adds -**

Factors influencing modeling utilization broadly reflected those for other research types, but findings provided more detail on influences for resource allocation policy decisions. This included the role of international funding organizations, the importance of flexible financing decisions, and the need for models to better align to country needs and decision-making processes, such as through integrated disease modeling. The role and means of stakeholder engagement was highlighted, and utilizing workshops can facilitate effective collaboration and communication between modelers and stakeholders and empower relevant stakeholders to interpret and apply modeling evidence.

**How this study might affect research, practice or policy -**

These findings may support modeling groups, sponsors, and stakeholders to collaborate, implement and apply modeling effectively to decision making for resource allocation. This study highlights opportunities to strengthen the local relevance, acceptance and utility of such analyses for better prioritized spending and ultimately to contribute to improved health outcomes.

## Introduction

Although countries have made critical gains in reaching global targets towards human immunodeficiency virus (HIV) and tuberculosis (TB) elimination,[1] closing the remaining gap in prevention and treatment targets will require strategic investment.[2, 3] As international HIV investment becomes increasingly constrained, countries are shifting to internal resources to fund their HIV responses. In low-and middle-income countries, the domestic share of HIV funding has increased from 51% in 2010 to 61% in 2020, and global funds available for HIV have been decreasing since 2017.[4] Similarly for TB, financing gaps to meet end-TB targets have widened since 2016, particularly among low-and-middle income countries.[2, 5, 6] Shrinking budget envelopes mean that governments need to make challenging decisions on HIV and TB investments amidst competing health and social priorities.

Mathematical modeling has been widely used to project epidemics at national and global levels, with models such as the Spectrum suite used to produce HIV estimates to guide national policy and planning.[7] Modeling is also recognized as an important tool when there are gaps in empirical data, or to compare the potential impact of different intervention combinations within a theoretical framework. The use of modeling to guide health policy decisions for infectious diseases has burgeoned in recent years.[8, 9] As well as projecting epidemics under different intervention scenarios, models can be used to inform financial resource allocation. Optima HIV and Optima TB are allocative efficiency models designed to do this.

Optima HIV and TB are dynamic, compartmental population-based models of disease transmission and progression for HIV and TB, respectively, integrated with an economic and program analysis framework. The models apply a mathematical algorithm to estimate the most cost-effective allocation of resources across disease-specific interventions to maximize progress towards specific objectives or country targets.[10–12] The projected impact that different funding allocations could have on epidemic outcomes is estimated and compared to outcomes if current spending patterns were maintained. The underlying epidemiology in each model application is calibrated to align with prior nationally-accepted estimates, and model outputs are validated with in-country stakeholders. Analyses are completed in partnership with national stakeholders and coordinating or funding agencies. The technical modeling team provide training of varying intensity to partnering stakeholders on using and interpreting the Optima models.[13, 14] Optima HIV and TB models have been applied in over 50 countries since 2010 and 2016, respectively, in collaboration with the World Bank, international and country partners. However, it has not yet been qualitatively evaluated how these models have influenced policy or financing.

Optima modeling studies produce research and economic evidence with the objective to inform health policy and financial decision-making.[15] The importance of evidence-informed policy has been highlighted by international bodies and institutions such as the World Health Organization (WHO) and National Institute for Health and Clinical Excellence, emerging from the paradigm of ‘evidence-based medicine’.[16–19] In practice, evidence-based policy is difficult to accomplish and often policy does not reflect the available evidence.[16, 18] Preferred terminology has shifted to evidence-informed policy, which moves away from assumptions of rational and linear decision-making and acknowledges that policy is developed in complex systems influenced by judgment, values, and social, political and economic factors.[15–17] A broad array of evaluation studies and conceptual frameworks, reviewed elsewhere, describe the utilization and impacts of research evidence on health policy.[18–22] Specific to modeling, examples include antiretroviral treatment guidelines being informed through modeling evidence and more recently, the use of modeling in the construction and adoption of elimination targets.[23, 24]

Resource allocation is critical to HIV and TB program outcomes, and improved targeting of resources can lead to substantial reductions in epidemic burden.[11] Despite the widespread use of mathematical modeling in health planning and policy, there are limited evaluations of how modeling findings and recommendations are adopted by countries into policy and programming, particularly in terms of resource allocation.[25–28] Although existing research prescribes stakeholder engagement as one of the key principles for effective modeling and researcher-stakeholder partnerships are a facilitator of research utilization in policy,[18, 21, 29] most modeling guidelines focus on technical aspects or results communication.[30] There is relatively less emphasis on how to work together with policy and decision makers.[31] This evaluation sought to assess the outcomes and factors influencing utilization of modeling research in health policy and practice from the perspective of the major consumers of Optima HIV and TB modeling: country stakeholders and their international partners. These findings will provide constructive insights on ways to improve the conduct and management of modeling research to maximize the relevance, perceived usefulness, and application of modeling results for setting health financing priorities.

## Methods

### Study overview

This study draws on qualitative data collected as part of an internal feedback and quality assurance (QA) exercise of Optima HIV and TB applications. In October to December 2021, the World Bank and Burnet Institute invited key stakeholders from countries involved in prior Optima applications to participate in a virtual interview to discuss their experiences and satisfaction with the modeling and engagement processes, how the model findings and recommendations had been used in country, and perceptions on the usefulness and impact of Optima analysis for supporting decision making and addressing evidence gaps. Qualitative enquiry through small group interviews was supplemented with quantitative case studies. These used more recent data to historically project the epidemic and estimate the impact of changes in spending or program coverage following Optima analyses. This analysis focuses on the qualitative findings.

### Participant selection and description

Study participants were purposively sampled from country stakeholders and international partner organizations who have participated in or funded Optima HIV and TB analyses. The research team compiled a list of all previous Optima HIV and TB analyses from 2012 until 2021. Any analyses using model versions prior to 2012 were excluded due to an inability to update the quantitative analysis, and analyses conducted after 2019 were excluded due to insufficient time elapsed to evaluate impact. The research team contacted stakeholders from 18 countries via email, purposively selected based on the above criteria and to maximize geographical region representation, to invite them to participate in an interview. Participation was voluntary. Up to three email contacts were made to encourage stakeholders to share their experiences with Optima. Where possible, representatives from a variety of organizations were invited and included, such as National AIDS Programs (NAP) or local equivalent, National TB Programs (NTP), Ministry of Health (MOH), the Global Fund and the Joint United Nations Programme on HIV/AIDS (UNAIDS).

Overall 54 country and international stakeholders working in nine countries agreed to participate in the qualitative assessment, representing a total of 11 country-disease contexts: five countries with an HIV analysis, four countries with a TB analysis, and two countries in which both HIV and TB analyses were conducted (Table 1). Two regional representatives were interviewed for their perspectives on multiple Optima HIV analyses conducted in their respective regions.

Within their organizations, participants’ positions included those relating to: strategic information, data analysts, monitoring and evaluation; program officers and specialists; program directors; planning and coordination; finance specialists; and specific to international health and funding organizations, country managers.

**Table 1.** Overview of study participants.

| Country / Region | Disease focus | Number of interviews | Number and type of interview participants |  | Language of interview(s) |
| --- | --- | --- | --- | --- | --- |
| Belarus | TB | 2 | 5 | NTP | English/ Russian<sup>*</sup> |
|  | HIV | 1 | 4 | UNAIDS, GF | Russian<sup>*</sup> |
| Botswana | HIV | 1 | 3 | NAP | English |
| Eastern Europe and Central Asia | HIV | 1 | 1 | GF | English |
| Eswatini | HIV | 1 | 5 | NAP | English |
| Kyrgyzstan | HIV | 1 | 2 | NAP, UNAIDS | English/ Russian<sup>*</sup> |
| Latin American and Caribbean | HIV | 1 | 1 | GF | English |
| Malawi | HIV | 2 | 1 | MOH<sup>‡</sup> | English |
|  |  |  | 1 | GF | English |
|  | TB | 1 | 4 | NTP | English |
| Mozambique | TB | 1 | 5 | MOH | English/ Portuguese<sup>*</sup> |
| Peru | TB | 1 | 8 | MOH | Spanish<sup>†</sup> |
| Ukraine | HIV | 2 | 2 | MOH | English/ Ukrainian<sup>*</sup> |
|  |  |  | 2 | UNAIDS | English |
| Zimbabwe | HIV | 1 | 10 | MOH, NAP | English |
| GF-Global Fund; MOH-Ministry of Health; NAP-National AIDS Program; NTP-National TB Program (or local equivalent) |  |  |  |  |  |
| <sup>*</sup> Interpreter present; <sup>†</sup> Facilitated in Spanish and notes translated; <sup>‡</sup> Technical consultant |  |  |  |  |  |

For the purpose of reporting confidentiality, study participants are only identified by stakeholder group (CS: country stakeholder; IFHO: International funding or health organization representative) to ensure confidentiality.

### Data collection procedures

Overall, 16 in-depth small group interviews were held with 45 country stakeholders and nine funding organization and NGO representatives (Table 1). Burnet Institute and World Bank representatives facilitated virtual interviews via the videoconferencing software Zoom using a semi-structured interview guide (see Supplementary Material). This provided flexibility to explore new themes as they arose while probing participants’ roles and satisfaction with Optima analyses and how model findings have had been used in country and impacted funding. Each interview involved between 1 and 10 participants and went for 45-60 minutes. If more key stakeholders were identified or were unavailable in the first interview, a second interview was conducted. Interviews were conducted in English with the exception of one led in Spanish. An interpreter assisted in interviews where translation was required. Interviews were audio recorded with verbal consent from participants.

### Data analysis

Qualitative data consisted of detailed and analytical notes taken during all interviews supplemented by partial transcriptions. Interview recordings were partially transcribed focusing on informational content except in three interviews impacted by recording error. Qualitative data were coded, sorted, stored and retrieved using Microsoft Excel. The Framework Method was utilized, applying an inductive approach to coding to allow for themes reflecting locally-situated and unexpected responses rather than embedding the coding in any single conceptual framework.[32] Following familiarization with analytical notes and audio recordings, the lead author used open coding to classify all data. Codes were grouped into broader categories based on similar or interrelated ideas and concepts concerning either the utilization or outcomes of Optima research, forming an analytical framework. The framework was iteratively refined in the process of comparing and contrasting emerging concepts, reducing the data for clarity, and until no new codes emerged. The resulting framework was applied onto existing categories and codes, charting a matrix of data by respondent group and codes. The interpretation process involved assessing the thematic data for characteristics, patterns and differences, exploring relationships between research utilization and impact, and interrogating the analytical framework against established theoretical frameworks of research use and impact.[18] Outcomes of Optima research were applied to an existing framework categorizing primary outputs, secondary outputs, and final outcomes.[18]

The coding and analysis was led by the lead author with input and review of the analytical framework from other authors during regular meetings. The results presented in this manuscript primarily focus on the factors influencing modeling evidence being utilized in health policy and practice.

### Public involvement

Participants were not involved in the evaluation design or conduct but were involved in the implementation of the original Optima modeling studies, including priority setting, data collation, result validation and interpretation. A draft version of this manuscript was disseminated to all study participants prior to submission with an opportunity to comment on how their experiences were represented or opt out of having their data included in the published manuscript. No participants objected to publication. The evaluation findings are also being circulated to participating groups as country case studies and a technical report. The findings of this evaluation will inform public involvement in future Optima modeling studies.

## Results

### Factors influencing research use

Six major themes were identified that act as facilitators or barriers to utilizing modeling findings in policy and financing decisions (Table 2). These factors are outlined in the sections below, describing the circumstances that aid or hinder modeling utilization.

**Table 2.**
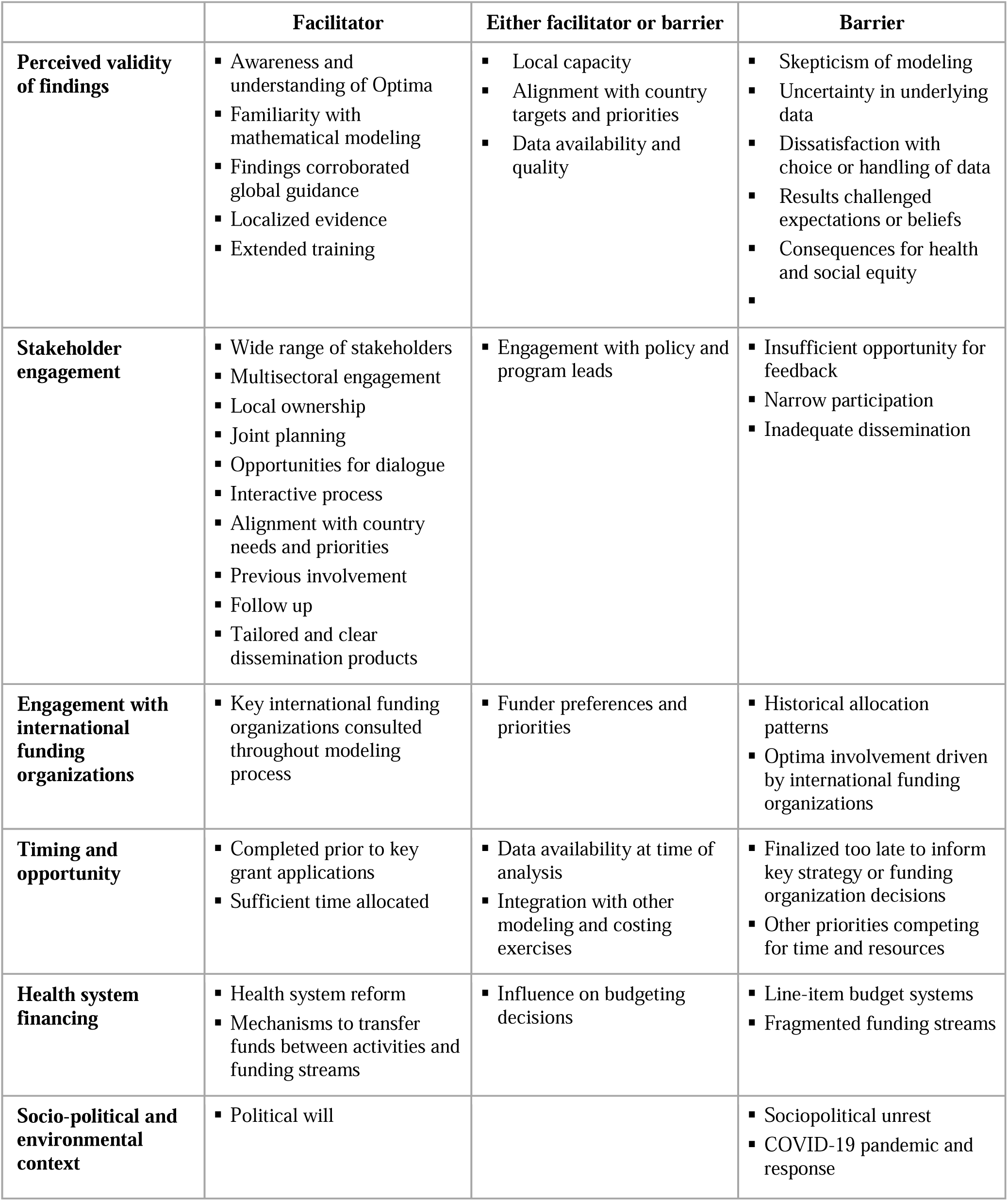
Facilitators and barriers of utilizing modeling evidence in policy identified in the evaluation.

## 1. Perceived validity of findings

Country stakeholders’ **awareness and understanding of Optima** and allocative efficiency impacted their perceived validity of Optima findings, which subsequently influenced decision making. Stakeholder groups demonstrating good awareness of Optima and allocative efficiency tended to portray broader acceptance and consideration of Optima recommendations in local policy compared to those with lower understanding of Optima modeling.

Participants also highlighted that the awareness and understanding of allocative efficiency among international funding partners was a determinant of research uptake and utilization, with interest and engagement in allocative efficiency varying between partners, teams and regions.

> “But I also have to say even with the [organization name removed], the topic of allocative efficiency is not as well spread as it should be.”(IFHO)
>
> “Besides people that work on it closely, I don’t think there was a lot of discussion about [allocative efficiency] … Of course we share the outcomes. People are aware of it. But it’s not about that. It’s about taking it and saying ‘OK, so this is the situation; what do we do now?’ How to push it. How to push the agenda with the information you gave us.” (IFHO)

Countries which had been involved in previous Optima analyses or other HIV Investment Case projects portrayed greater confidence and knowledge of Optima methodology and capabilities. In these cases, **familiarity with Optima and allocative efficiency** had facilitated more meaningful engagement with the processes, including the critical assessment of data inputs and findings, and enabled greater ownership.

> “However the concept of efficiency, I would say that one we are familiar with for we have done some investment case on HIV prior. And in essence investment case is trying to help guide on the level of investment that can have an impact for benefits. And often on the long-time horizon, which is what Optima has also talked to.”(CS1)

Perceived validity of findings was also influenced by accordance of findings with expectations, organizational beliefs and alternative evidence. Model outcomes that confirmed existing assumptions or other evidence could be used to justify spending resources and strengthen funding applications.

> “There were no surprising findings, but it validates the intuitive thinking.” (IFHO)

Where **findings corroborated global guidance** or institutional beliefs, Optima added to a local evidence base and increased motivation for utilizing the findings in resource allocation. For instance, in one country Optima findings provided contextualized arguments to support the shift to universal antiretroviral therapy (ART) access, in line with World Health Organization (WHO) guidance, leading to increases in investment in ART.

> “The finding of the Optima exercise has strengthened that thinking and that expectation.” (CS4)

In another example, Optima findings were actively used in discussion with government bodies to scale-up opioid substitution therapy (OST), helping to overcome legislative barriers to OST prescription and gain support for OST both within and external to health departments. Despite there being considerable existing evidence supporting interventions such as OST or universal ART, stakeholders valued the ability of Optima to provide **localized evidence**.

Conversely, in some settings where **results challenged expectations or beliefs**, the modeling evidence was discounted or not considered for financial decision making. This was often connected to data availability or population size uncertainty, and in some applications the reliance on assumptions or limited data negatively affected the perceived validity of the results.

Despite commonly reporting involvement in data collation, in select cases respondents communicated **dissatisfaction with the choice or handling of data** used in the model. Perceived validity was also compromised in select examples when there were discrepancies between Optima HIV and epidemiological projections from other HIV epidemiological models used for annual planning and forecasting.

**Alignment with country targets and priorities** was also an important consideration. In one instance, respondents conveyed that pre-exposure prophylaxis (PrEP) was being scaled up, despite this not being supported by model findings, due to preestablished funding and government priorities. Diverse stakeholders, including program planners, policymakers and donors, emphasized that the policy environment is increasingly structured around cross-cutting programs, including structural interventions and integrated disease programs. Respondents portrayed that reflecting these elements in modelling would facilitate broader utilization in policy decisions.

In some settings respondents conveyed that there was a general **skepticism of modeling** among some personnel involved or key decision-makers. This was not unique to Optima but had led to some stakeholders being doubtful and critical of modeling evidence. They attributed this to doubts regarding assumptions and concerns that the process of model calibration is overly subjective and prone to biases. One country representative explained that evidence generated through modeling was perceived as inferior to empirical data derived from programs or studies.

> “When they see the back of it and appreciate that they can play with the figures, then it becomes more [of a] political instrument rather than [an] epidemiological instrument.” (IFHO)

In one setting with pronounced **uncertainty** on population size estimates, stakeholders believed that the proportion of new HIV infections among people who inject drugs (PWID) had been overestimated in the Optima model and thus the recommended scale-up of PWID prevention interventions was not justified.

> “The last two rounds surfaced a negative factor for [country]: 50 percent of the new HIV cases come from IDUs, which in reality is not the case. We did send our remarks emphasizing that we do not have such a high rate among IDUs.” (CS8)

A few means were discussed to strengthen knowledge and awareness of Optima and modeling processes. Most eminently, **extended training** and building local capacity to use Optima were portrayed as a key route to developing local ownership.

> “Need to increase the ownership of Optima here locally through a greater amount of trainings. Yes, we have gone through the five-day long workshop, but it was quite intensive yet still not enough. We need more hands-on training later on. Optima math modeling is quite sophisticated, and not something you will get fully in five days. And what we need essentially here is the experts or resources who would be capable of using Optima on their own, to the extent possible. So more training would be required.”(CS2)

Although the training provided from Optima was appreciated in many settings, respondents from one country conveyed that no training had been received, and respondents from multiple countries suggested that more training would have been useful, particularly for technical leads. Extended training may build local skills and capacity to support, interpret and independently utilize modelling studies.

> “In terms of training, I think it was useful. For each country group there was a representative delegated by your institution, I believe. I don’t remember, to be honest, who cooperated with us, but it was quite interesting and efficient.” (CS8)

Based on input from funding organization representatives, conducting training and modeling as part of a collaborative workshop may offer a more immersive experience for participants and facilitate meaningful involvement, dialogue and collaboration.

In some settings, recommendations to deprioritize interventions serving specific population groups or with social and health impacts outside of HIV/TB were reportedly met with resistance among stakeholder groups who represent the needs and service delivery among implicated populations. In these cases, certain modeling outputs may have **consequences for health and social equity**. This lowered the perceived validity of findings among some stakeholders.

> “We faced some challenges and resistance while reallocating those funds, specifically on the side of civil societies and NGOs because due to the Optima results, we had to taper off funding from one community and groups towards the others.” (CS2)

## 2. Stakeholder engagement

### Stakeholder selection

The type of stakeholders included in Optima analyses varied by country, but generally involved representatives from the NAP, NTP and/or MOH, as well as potentially NGOs and civil society organizations (CSO), funders, and international organizations. Country partners were encouraged to involve all perceived relevant stakeholders. Country respondents generally appreciated engagement of a **wide range of stakeholders** representing different categories – from technical to policymakers – and including multisectoral representation.

> “We had a workshop; two workshops with other stakeholders and the team of Optima also. … And in these workshops [other stakeholders, NGOs that work with us] also were invited and we discussed some data with them. They were involved in the process. And some data also that MOH didn’t collect we also received some of them from one of our partners. It was good involvement of all stakeholders. Even for dissemination of results also we shared with them some of these results.” (CS6)
>
> “The joint oversight committee meeting is the one that is critical because the stakeholders, or the members rather, are the CEOs and country directors of all of the stakeholders in the HIV/AIDS response. And therefore those are the decision makers of allocation of funds.”(CS1)

Funding organization and international stakeholder representatives perceived that effective engagement with in-country stakeholders, and particularly the MOH, was integral to fostering **local ownership** of Optima products.

> “This component of ownership is probably the hardest, and it’s the one that was emphasized a lot. It started with a letter to the Ministry of Health from us. And then it was, when you own it, you designate that this is your report. And then they validate the reports, and so on. And some cases it took quite a long time to validate the final reports, but we got to it, you know. Despite all of the other priorities that were running. So in that sense, I don’t think that any country will tell you ‘they don’t know’, ‘we didn’t do it’, ‘it’s something that [IFHO] did’. It’s not the [IFHO], they did it.” (IFHO)

In one setting, respondents attributed the lack of ownership to minimal follow up, discussion and influence of Optima results on policy.

> “It’s an important factor, that the [name of institution removed] which was the key institution, to own the results of this analysis-modeling exercise. The [Institution] did not really promote Optima. Has not brought up Optima recommendations in discussions about the Global Fund Proposal. Or the PEPFAR funding. Or the National AIDS Strategy. I still think they have not released this document as final.” (IFHO)

### Types of participation

Country representatives most commonly reported being engaged in **data collation**. There were fewer examples of country teams that had been directly involved in model calibration and projections. However, country teams that had participated in repeat analyses expressed enthusiasm and knowledge to be more involved in future analyses.

“Speaking of the NGOs and civil societies, their role was limited to providing the data. How this data was interpreted and how the reallocations were contemplated later on …were handled by the Optima team, by the people from Geneva UNAIDS, by the Optima taskforce. I don’t think that the civil societies and NGOs contributed specifically to this technical work.“ (CS2)

Even at the level of data collation, joint participation between technical partners, country stakeholders and IFHO was perceived to help foster ownership, acknowledgement of study validity, and the actioning of study recommendations.

> “It’s not that we say ‘you should invest more in treatment because we are the [IFHO]; We say, ‘we together ran the analysis, here is the science behind it. This is how much money we need to hit the 90-90-90…It helps to have, you know, ‘we’ve done this together, this is what the data says, so this is probably where we should move’.” (IFHO)

Participants valued meaningful **opportunities for dialogue**, and in isolated cases respondents recalled insufficient opportunity to provide feedback to their country leads, thus limiting the perceived validity and subsequent application of findings.

> “I think it’s a matter of discussion and preliminary work with national team – discussion and finding a common ground.” (IFHO)
>
> “We were not able to follow through the whole process because we were not in a position to give feedback to our principles [country leads].” (CS3)

Dialogue and **follow-up** both within funding organizations and between funding organizations and country partners was also viewed as a critical determinant of allocative efficiencies being applied and used for funding decisions.

> “Since I arrived, I don’t see anything happening with the outcomes of the Optima study. The idea was, that this: the Optima recommendations would then translate in in the next funding cycle of the Government. From what I know, and I tell you only from a [organization name removed] employee working in a very limited area – I was not part of any conversation or follow up of that. And I don’t think it happened. So for me it’s a wasted opportunity.” (IFHO)

In numerous settings, participation was enhanced through **joint planning** between country partners and the Optima team. Effective joint planning supported **alignment with country needs** by enabling tailored analyses adapted to specific country priorities and required budgeting decisions, such as district-level analyses to support geographical targeting of voluntary male medical circumcision or analyses focusing on the impact of COVID-19 service disruptions.

> “It was the NTP that proposed the modeling exercise should be done at district level.” (CS4)

Other characteristics of **effective participation** described by respondents included interactive processes, opportunity to review data inputs and outputs, technical assistance from the Optima team, and consistent follow-up.

### Stages of involvement

Interview respondents described ways to strengthen participation by engaging stakeholder groups in different ways over the course of Optima study implementation. Engagement of a wide range of stakeholders was considered most important during study set-up and dissemination to facilitate awareness raising, information sharing, contribution to research questions and validation of findings. Multisectoral involvement including NGOs and international stakeholders was perceived as particularly important during **dissemination** to support policymaker and funder buy-in. For instance, representatives from one participating country emphasized the importance of discussing findings with ministries outside of health to gain broader support for interventions such OST and to influence national strategy. Four settings reported on inadequate dissemination, due to either language, form of dissemination materials or insufficient sharing of dissemination products.

> “There was a gap in dissemination and the evidence that has been generated did not get to the critical programmers who make the decisions in these programs.” (CS3)

Aside from stakeholder engagement, voiced preferences for dissemination included tailored briefs for sub-groups of stakeholders, hard copies of briefs and reports, translation into local language, and concise and simple language.

Respondents from some settings suggested that **earlier engagement** of some stakeholders, such as funding organization and CSO representatives, could facilitate understanding and support of Optima findings.

> “It would be beneficial to include someone from civil society in this taskforce. If excluded, they might think the government is hiding something or has some sort of hidden agenda. We will gain more support and understanding from them if that person will be included.” (CS2)

Nevertheless, there were differing views on whether **policy and program leads**, including government officials, should be involved earlier and throughout the analysis to promote ownership of findings, or whether engagement with these leads was more useful when restricted to stages such as defining national policy questions and dissemination. Funding organization representatives valued focusing participation on monitoring and evaluation (M&E) personnel to promote Optima as a data-driven technical exercise.

> “So the way we organized this we didn’t do the political part. We sold it as a very technical exercise. And we involved people from the M&E departments. There were no decision makers, just the people who do the data, the M&E. So the result was it was a very helpful exercise…” (IFHO)

**Previous involvement** with Optima facilitated stakeholder engagement and increased demand for future modeling, thus may have facilitated utilization of modeling evidence.

> “Optima was quite a valuable tool, not only from the standpoint of empirical or fiscal data, it also provided a lot of practical oversight for both the Government, Ministry and the civil sector in terms of how money was spent, where we should go, what results we would get. Especially it was the case in the second time. Because we implemented this Optima model a second time now.” (IFHO)
>
> “We are now aware of how the model works; it would be good to maintain a working relationship.” (CS1)

## 3. Funding organization influence

Respondents from several settings identified that **funder preferences** had a bigger influence on resource allocation than Optima findings, although the two were not necessarily exclusionary. In some cases, funder support may have reinforced the scale-up of interventions such as HIV self-testing, which was prioritized in several Optima analyses. In a couple of cases respondents perceived that funders discounted the research findings to comply with their own priorities.

The extent of **funding organization involvement** in Optima analyses was also a determining factor for research utilization. Some funding organization representatives acknowledged that where funders exclusively drove Optima implementation, it may have lessened country ownership and reduced the analysis to a requirement for funding applications rather than a tool for decision making. In contrast, in some settings respondents felt that involving all key funders earlier in the Optima process could better support the use of findings for resource allocation decisions. Limiting funding organization involvement to a single sitting for dissemination of findings may have inhibited the appreciation and uptake of findings, particularly in settings receiving international funding from multiple entities.

> “They were hearing [of Optima analysis] for the first time, and being on the tail end of it. Which might have been a little too much to chew at one sitting…It would work better if we get them through the mill earlier on before we get to the results.”(CS1)

Country representatives and funding organization representatives both relayed that budgets tended to favor **historical allocation patterns**, which limited depiction of research findings in policy and financing decisions in some settings.

> “Funding for subsequent years would usually be predominantly based on the previous year. So it’s more of a recurrent budget and budget proportions, not necessarily guided by evidence of impact.” (CS1)

## 4. Timing and opportunity

The timing of Optima analyses was a key determinant in application of findings. Analyses and dissemination that were **completed prior to key grant applications** or strategic development had opportunity to inform these decision processes. In some cases, strategic planning could still be informed by preliminary findings from the modeling.

> “The exercise was still being done during the strategic plan development and also the NFM [New Funding Mechanism] grant making. I think this was the major times when there has been key decision making and resource allocation. Since we had some preliminary output from Optima, I think those recommendations were also being considered and incorporated in those kind of exercises.” (CS4)

In other examples, due to delays in model implementation or stakeholder review, unforeseen circumstances, as well as different planning cycles, analyses were **finalized too late to inform key strategy or funding organization decisions**. Both country and funding organization representatives spoke about the importance of factoring in sufficient time for preparation, country engagement and data collation.

Timing was also mentioned in relation to **data availability**. Delays in analyses may be caused by waiting for new data, such as updated National AIDS Assessments (NASA) reports. Interviewees found that the timing of analyses should be intentionally aligned to utilize the most recent epidemiological or financial studies.

> “The full analysis didn’t account for these [latest] prevalence survey results, because we just had the results last year. But Optima was conducted with the data to 2017.”(CS6)

**Competing demands** at the time of Optima analyses was a limiting factor for stakeholder engagement. In one example, the Optima application happened simultaneously with the development of the M&E framework for the new National Strategic Plan, thus limiting staff availability to support the Optima analysis. More broadly, funding organization representatives noted that competing priorities and busy agendas, particularly in the context of the COVID-19 pandemic, made it harder for country stakeholders to engage with Optima and had delayed several analyses. In some cases, stakeholders recognized that competing priorities coupled with limited resources increased the perceived value and application of allocative efficiency analyses.

> “The issue of allocative efficiency always remains a priority in terms of our response because we don’t have adequate resources and we also do have now quite a number of competing priorities.” (CS3)

Representatives from one participating country raised poor timing of analytical activities due to **inadequate integration of various modeling and costing exercises**. This led to a duplication of effort to track resources and collate data, such as for NASA and National Health Accounts, and Optima and Spectrum.

## 5. Health system financing mechanisms

In some settings budget reallocations were constrained by the health system financing model. Line-item payment systems define the total amount to be financed to a hospital or other organization based on the expected costs of clinical and non-clinical staff, equipment, medicines, utilities, and maintenance, which are determined based on expenditure in the previous year.[33] Systems that utilized **line-item budgets** to finance in-patient services lacked the flexibility and incentive to transition to ambulatory, decentralized care models, even when Optima findings demonstrated alternatives to be more cost-effective and efficient. Without changing how health services are funded at a structural level, adopting these changes would lead to reductions in hospital funding in subsequent years. In these cases, the system lacked appropriate **mechanisms to transfer funds**, such that savings could not be reinvested in alternative activities nor for comparable care outside of the hospital setting.

> “The number of beds have decreased, but this does not mean that they release the funds … for other activities.” (CS5)

**Fragmented funding** of epidemic responses also posed challenges to redistribute either existing funding or cost-savings through implementation efficiencies. Respondents from several settings spoke of difficulties in transferring cost savings from general population HIV testing programs to key population prevention and testing due to different sources of program budgets.

> “Of course, for [country name removed] Government it is easier to fund HIV treatment and expenditures related to treatment, care and support for HIV positive people. The Government covers around 80 per cent of such expenditures…In terms of the prevention programs … the cost sharing in this case is the opposite: 80 per cent will come from Global Fund.”(CS7)

In one case this meant that while the state budget increased commitments for key population prevention in line with Optima recommendations, eventually these funds were not disbursed due to the complexity in actualizing resource reallocation. Subsequently, **health system reform** was identified as a key enabler of resource allocation in line with cost-effectiveness considerations and specific Optima recommendations.

In one setting, health system reform was underway to reduce the dependency on donor funding, promote sustainability and improve the efficiency and quality of care. This included the introduction of social contracting, enabling public health centers to purchase HIV prevention services through non-government and CSOs. In this example, the respondent felt that making the government the main recipient of international funding gave them more power to advocate to local governments and increase funding for key population programs through social contracting, in line with modeling recommendations. They described this as a lengthy process taking place over two-to-three years which eventually enabled better adoption of allocative efficiency findings.

> “So, [local governments] begin to see the benefits of these expenditures now as well as the purpose of it. Thus it is much easier to convince them now that funding for these areas should be increased. So, the role of the main recipient as a main driving force in advocating state social contracting is very strong.” (CS7)

Optima commonly engages most closely with the NAP and NTP for HIV and TB studies, respectively. Some respondents referred to these parties having **limited influence on budget** decision making. Their actions were constrained to making recommendations to the relevant government structure, but this was insufficient to enable the application of Optima findings to resource allocation in some settings.

> “The NTP is not the owner of the government funds. It has limited influence on how the funds are allocated.” (CS5)

The impact of **indirect spending** such as management costs and overheads for the national HIV program cannot be readily quantified in terms of direct impact on defined disease transmission parameters and are usually considered as a fixed expense in Optima allocative efficiency analyses. In practice, country teams often found it difficult to differentiate direct from indirect spending. Some respondents reported a need to better understand indirect spending, such as administrative costs and supply chain management, to inform resource allocation.

> “The performance of a program depends on various factors, which I believe need to be included in future modeling to bring robust results…For instance, we have a shortage of human resources – it’s one of the determinants of the performance.” (CS6)

## 6. Socio-political and environmental context

Respondents considered that **political will** was important when findings concerned key population groups or migrants, which were politically sensitive in some contexts. In settings where related behaviors are criminalized or these groups are not prioritized in existing resource allocation, Optima findings were more likely to face resistance from policymakers, particularly outside of the NAP or NTP or at a sub-national level. This was particularly pertinent in settings in the process of **transitioning from international to domestic spending**.

> “A governor in a region may have never heard before how MSM [men who have sex with men] are influencing pandemics and now he is told that funds need to be allocated for condoms for this group, for lubricants. Usually, these things come as a shock to local government.” (CS7)

Despite the expected resistance in these settings, funders perceived that Optima supported advocacy by providing objective rationale for prevention programs for key populations. Optima provides an opportunity to “put the science before the politics” (IFHO), and in this way has the potential to depoliticize resource allocation.

External factors such as **socio-political** unrest and the COVID-19 pandemic also impacted the influence of Optima findings on resource allocation. In one setting, conflict and the existence of autonomous regions meant that sub-national HIV programs and services are not controlled or funded by the Government, and thus not influenced by Government resource reallocations. The **COVID-19 pandemic** placed strain on healthcare resources and budgets, necessitating diversion of funds from other health programs.

> “Every free cent was reallocated to COVID.” (CS8)
>
> “In the context of [country name removed] …there are lots of conflicting priorities. … The concept of efficiency was not high on the agenda. The other problems they are facing, in their view, are more important. No human capacity or time to focus.” (IFHO)

In some settings, respondents reported increases in service costs due to COVID-19, further impacting the relevance of prior allocative efficiency analyses.

> “Especially since COVID, as these costs have increased since then. Now you have to go to three different health care institutions to have your diagnosis confirmed and have treatment allocated, which is laborious. If there was one single location, these costs could be reduced.” (CS7)

### Outcomes of Optima applications

Country stakeholders and funding organization representatives conveyed that Optima HIV and TB analyses have led to a range of outcomes which can be broadly grouped as primary outputs, secondary outputs, and final outcomes.[18] These correspond to (1) outcomes improving knowledge and benefiting future research; (2) reflection of modeling evidence in planning, decisions and advocacy and improved allocation of resources; and (3) gains in cost-efficiency or funding, summarized in Figure 1 .

**Figure 1.**
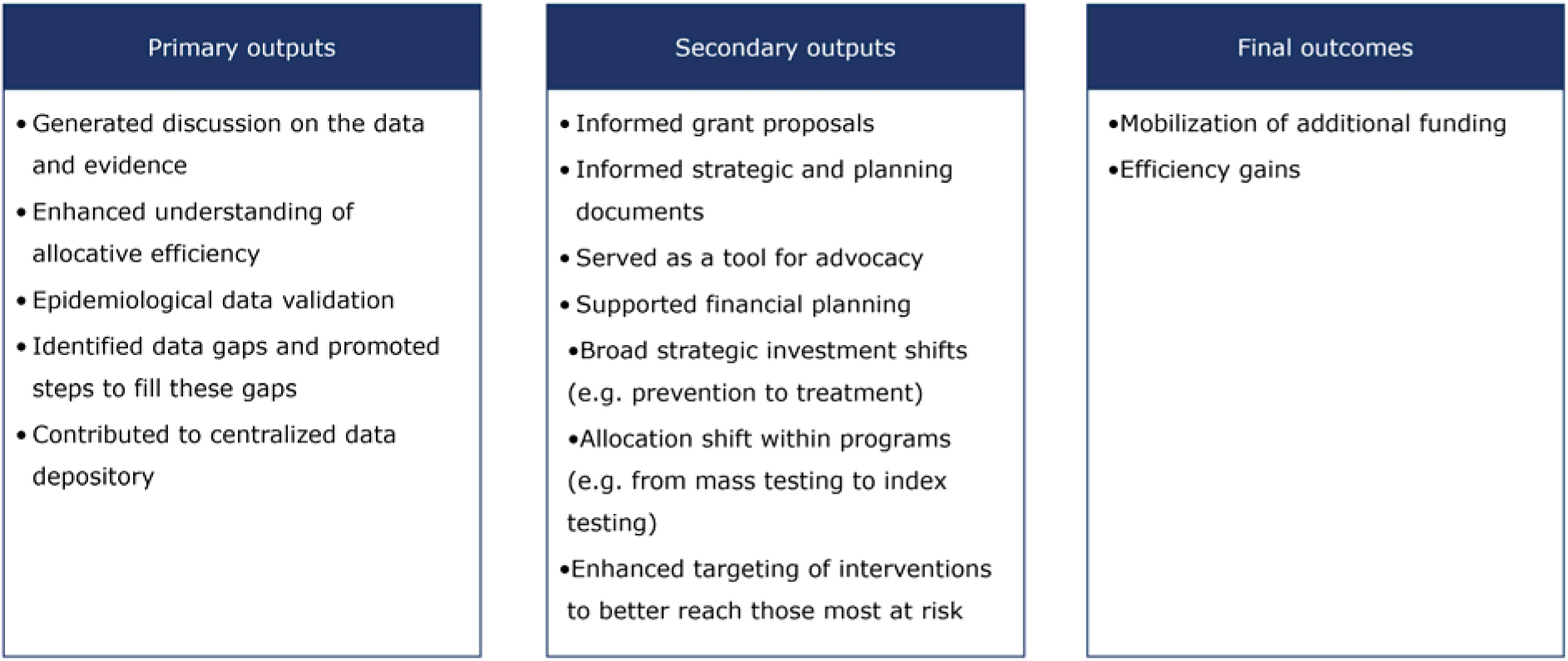
Overview of outcomes reported from involvement in Optima HIV and TB analyses.

Although final outcomes have greater significance to cost-effective programming and ability to impact epidemic measures, many respondents also acknowledged the value of the more immediate primary outputs such as generating internal dialogue and strategic thinking on programs, priority populations, and data gaps.

> “The exchange of the results of this modeling – what works, what doesn’t work, what data we have…catalyzes internal discussions about the quality of data and the completeness of data.” (IFHO)

The most common application of Optima HIV and TB analyses reported by interview respondents was to inform grant proposals and strategy documents. The timing of Optima analyses was often tied to Global Fund funding requests, and Global Fund grants were most frequently cited as having been informed by Optima. Specifically, respondents referred to using Optima findings to help set and review targets, select high-impact and cost-effective activities, and plan budgets. This was corroborated in publicly available grant proposals. Some respondents referred to using Optima findings to support advocacy for new investments, such as changes to modalities for testing and treatment delivery.

> “With Optima study, we can really see where we can negotiate or request funding. [By] showing evidence that by doing this and that intervention we can really have more impact and help the program in [country name removed].” (CS6)
>
> “As a country I think we did benefit in terms of the exercise because it opened our eyes in terms of the different programs where we need to prioritize as a country; where we can spend less and get more from” (CS9)

In four settings Optima-informed advocacy led to mobilization of additional funding.

> “One of [the Optima recommendations] was to expand the coverage of community outreach and preventive treatment among high risk and vulnerable populations. It indirectly oriented us to advocate for money. … so eventually that guided us to allocate more in these two particular interventions.” (CS6)

A representative from an IFHO described how Optima had provided supporting evidence for ongoing advocacy efforts to reduce the price of antiretrovirals, given differential pricing made obvious through a regional Optima exercise.

Funding organization representatives in particular considered that Optima provided an objective basis for policy advocacy, which was particularly useful when results supported increased funding for programs not readily supported by the government agenda, such as the scale up of OST.

> “The biggest breakthrough in our case was with opioid substitution therapy. In 2018 the expenditures were at US$ 103k, while in 2022 they committed US$ 800k with an increase to US$ 825K in 2024.” (CS7)

## Discussion

This evaluation confirmed there are a multitude of factors influencing the use of modeling to guide resource allocation for national HIV and TB programs. Allocative efficiency modeling has a supportive role in opening dialogue around budget allocations, providing localized evidence to support planning and advocate for funding, and helping policymakers visualize the benefits and consequences of different scenarios, including maintaining the status quo, to enable informed policy decisions. Facilitating stakeholder engagement and collaboration between policymakers and researchers can help to bridge modeling evidence, policy and practice for greater impact in achieving health outcomes.

Factors influencing the utilization of modelling evidence for administrative policy decisions closely resembled those described for other forms of research evidence.[19, 21, 34] In particular, perceived validity and relevance of modelling evidence, timeliness, appropriate dissemination of research, and strong collaboration between country stakeholders and research staff were key facilitators. These resonated with elements of multiple conceptual frameworks of research utilization in policy, including the RAPID Framework[22] and the Interfaces and Receptor Framework.[18] These frameworks acknowledge the complexity and multidirectional interactions between the research and broader political and funding environment, which may be navigated through interfaces or linkages between researchers and the users of research.

Modeling was considered less credible for informing policy decisions among select stakeholders due to assumptions and potential subjectivity. Although several studies recognize the importance of the type of research or evidence in affecting uptake for policy decision-making, modelling science is rarely explicitly mentioned in these texts.[16, 19, 35] These findings support the work of Rhodes and Lancaster (2020), which highlights that inherent uncertainty in modeling, use of projections and abstraction can mean they are viewed as a weaker form of evidence by some decision-makers.[9] A review of use of modelling evidence in WHO guidelines showed that modelling-informed guidelines were considered to have lower quality of evidence than guidelines not informed by modelling evidence, although this may be biased by when modelling evidence is preferentially used.[36] Strengthening collaborations between researchers and country stakeholders, particularly through participatory modeling, can improve the quality of models by maximizing available data to parameterize models,[37] and may thus help to promote confidence in modeling evidence. Several researchers have endorsed the idea of models as a “boundary object” which even through disagreement or uncertainty, can promote multidirectional dialogue and exchange to support priority setting and mediate knowledge into policy decisions.[9, 28, 37] In this way, the process of modeling can become as important as the outputs and research products.[37] However, findings that contradicted stakeholders’ beliefs were, at times, rejected in the current evaluation. Providing sufficient opportunity for diverse stakeholders to deliberate and discuss findings may help prevent disagreement from blocking evidence utilization.[9, 21, 25] This may be particularly important for programs that may be politically polarizing, such as those serving key populations.

In practice, interviewees described a wide spectrum of stakeholder participation in Optima analyses. The role of engagement with varied stakeholders throughout the research process for better policy decisions is highlighted particularly in network-based models of policy-making and participatory modeling approaches.[18, 37–40] We found that utilizing workshops can facilitate direct engagement, collaboration and effective communication between modelers and stakeholders and empower relevant stakeholders to use and interpret modeling. Findings also supported the value of previous involvement in modeling to build on local capacity and support local leadership in applying modeling to HIV and TB program decision making.[18] Further, repeat modeling, which was more common in the context of HIV, can drive data collection and availability. However, high turnover of personnel in some positions may potentially be a barrier to building institutional capacity and familiarity with modeling.[21] Where models are not institutionalized at a national level, additional funding may be needed to support renumeration and mobilize appropriate national consultants and teams to engage in the modeling process.

In general, research evidence can be used in policy in different ways, categorized as: instrumental, with explicit application to policy formulation; symbolic, whereby it is used to justify a position already taken; or conceptual, leading to a gradual shift in awareness, understanding and perceptions.[18, 20, 34] Although modeling was not always attributed with providing new evidence, this evaluation showed that contextualizing empirical evidence and global guidance through modeling can enhance acceptance and adoption of evidence-informed practices, demonstrating conceptual evidence use. Modeling findings were frequently cited as consistent with resource allocations designated within funding proposals and strategic planning. While some respondents supported the role of Optima in instrumentally informing these decisions, other research has reported on modeling evidence being used symbolically to support existing decisions and strategy.[18, 25, 34] International funding organizations, government bodies, and public health researchers may have preconceived notions or agendas, and evidence that resonates these assumptions and priorities is more likely to influence policy.[22]

Findings demonstrated higher perceived value of modeling among stakeholders when it was built around country priorities, timing of policy decision-making, and the broader health system context. Consistent with other studies, we found that international funding organizations often initiate the analyses or define the modeling objectives.[26] Funding organizations may have a more vertical approach than national disease or health programmes,[41] which may limit the perceived usefulness and national uptake of findings. Both national and international stakeholders conveyed a need for modeling to support integrated disease and program delivery, structural interventions, and decisions around human resources and program administration. By not accounting for these, disease-specific models may not fully capture operational feasibility and decision-making processes, thus undermining perceived validity.[42] Multi-disease, “whole system” approaches which consider interactions between diseases, interventions and the health system, such as for the Thanzi La Onse model in Malawi, may enhance the relevance of modeling for local decision-making and resource allocation.[43, 44] Further modeling which considers the workforce capacity and subsequent costs will facilitate transparent and feasible estimates of providing cost-effective interventions.

Research utilization in health policy relies on the recommendations being politically and administratively feasible.[35] The realization of resource allocation in line with allocative efficiency is dependent on the flexibility of funding within a health system, particularly whether funds can be reallocated between programs. Financing for key populations prevention programs is frequently still provided by international funders through CSOs, while domestic funding often covers care and treatment. The evaluation findings demonstrate that this fragmented investment approach was often a barrier to redistributing funding from generalized testing and prevention to programs focusing on key populations. Many countries dependent on external funding are graduating to domestic funding in the near future.[45] Mapping of the responsible funders and their recipients, expenditures, coverage, and consequential impact of all programs through modeling allows for an evidence-informed list of interventions that could be prioritized by local governments. Social contracting is one proposed mechanism to promote the strengths and sustainability of CSOs’ role in tailoring HIV and TB services towards the needs of those most vulnerable. It involves governments contracting CSOs to delivery services to key populations, thus channeling government financing for community-based outreach and care initiatives.[46] A 2021 study analyzing the sustainability of CSO provided services in the Eastern European and Central Asian region found that these services, often provided by peers for those most marginalized, are slow to be financed by local governments,[47] which was also demonstrated in this analysis. Collaborative modeling exercises can support the implementation of social contracting by engaging partners in constructive dialogue and consensus building on target setting and budget allocation.[48]

A number of limitations should be considered when interpreting these findings. The results may not represent the views of institutions and individuals who were non-responsive to multiple interview requests, and thus biased to settings with higher engagement in Optima processes or a more recent Optima analysis. High departmental staff turnover and secondment from HIV/TB to COVID-19 may have also impacted participation. Interviews were facilitated by the Optima research team with the potential to introduce social desirability bias. However, the interviewers encouraged honest feedback and participants relayed both positive and negative experiences and outcomes. This evaluation focused on country stakeholders and funding organizations as key consumers of Optima research, with limited input from international health organizations. Findings may not be generalizable to other entities utilizing modeling evidence, such as WHO and UNAIDS. Due to technical error, verbatim quotes were not possible to include from several interviews, and analysis relied on detailed notes. In some instances, quotes were conveyed through an interpreter and may not represent the exact wording of the original speaker.

In conclusion, this evaluation demonstrates that allocative efficiency modeling has supported evidence-informed decision making in numerous contexts, enhanced the conceptual and practical understanding of allocative efficiency and supported constructive dialogue on the data and evidence. Understanding factors that influence utilization of modeling evidence has relevance to diverse modeling groups informing health program and financial planning in international contexts. To facilitate the utilization of modeling results in health policy and practice, modeling collaborators should time analyses prior to key strategic and financial planning exercises, involve diverse stakeholders at key stages, and develop stakeholders’ confidence and understanding of modeling. To further improve relevance and acceptance of modeling findings, there needs to be greater consideration given to integrated disease modeling, equity goals, and financing constraints.

## Data Availability

Data from this evaluation is not available for sharing as per ethics agreements. Data and reports for underlying country models are published and openly available in the World Bank Open Knowledge Repository and through Optima.

https://openknowledge.worldbank.org/

http://optimamodel.com/

## Footnotes

## Acknowledgements

The Burnet Institute and the World Bank are grateful for all feedback provided by participants from country teams and from international partners and acknowledge the founding work of all personnel and collaborators who provided input on the original Optima analyses. The team extend their gratitude to Anna Roberts for support with project administration and data collection, to Jaime Nicolas Bayona Garcia, Roman Kulchynskyi and Felix Tsoy for providing translation support during interviews, and to Nisaa Wulan and Lung Vu for their contributions to the analyses and interpretation.

## Contributors

The evaluation was conceptualized and planned by NFH, NC, NS, DtB, and RMH. Data collection was carried out by DtB, NFH, ALB and RMH. Data analysis was led by AB with support from DtB. Manuscript writing was led by ALB. All authors contributed to the interpretation of the data, manuscript review and editing. NS is the guarantor. The corresponding author attests that all listed authors meet authorship criteria and that no others meeting the criteria have been omitted.

## Ethics approval

The original QA exercise was not subject to ethics review. The Alfred Hospital Ethics Review Committee approved the collation and analysis of the program QA data (project number:158/22). All study participants were notified of the use of their data for extended research purposes and had the opportunity to opt out of further research.

