## Supplemental file 1 for "Qualitative evaluation of the use of modelling in resource allocation decisions for HIV and TB"

**Interview Guide: Country Stakeholders**

*Intro Script/ recap of the study objectives and key findings/ recommendations*

**Background information of the stakeholder(s) being interviewed**

- Name (can be initials):
- Position:
- Involvement with the study:

**General study context**

1. What was the rationale as to why the study was conducted?
   - Prompt: expected reduction or increase of annual HIV incidence, funding application, to explore intervention efficiency improvements
2. Which key stakeholders were included (eg. Ministry of Health or National AIDS committee)?
3. How many individuals (approximately) participated actively in the Optima HIV/TB application?
   - Prompt: as providers of data/information, participants in the model application, reviewers of data inputs or results
4. Did the Optima implementation improve the understanding of the allocative efficiency (AE) concept and its applications?
   - Was there a training on AE and/or Optima?
   - Was the study led by the country team with technical assistance from the Optima team?

**Epidemiological and economic modelling outcomes and impacts**

5. What, in your opinion, were the most important messages from the Optima analysis?
6. What effect (if any) did the Optima application have?
   - Prompt: Supporting strategic thinking? Supporting decision-making on financing? Providing evidence for advocacy? Addressing data/evidence gaps? Guide planning?
   - Did the modelling results change the way you understand the epidemic?
   - Did the study results improve policy makers' understanding of epidemic trends (historical/projected)?
   - Do you have any feedback on the accuracy of the projections/ estimates (trusting, perceptions)?

7. What implementation efficiencies or cost-savings – if any – were found in-country following the study?
   - Are you aware of any changes in program implementation or in budget/ funding allocation (to sub-programs/ specific interventions) made in response to the Optima study?
   - Were the modelling results used to inform funding requests?
   - Were any follow-up studies done to investigate costs, coverage, or unit costs?

8. Could you please briefly describe to us the current HIV /TB landscaping (programming, financing, etc.) in the country?
   - Any changes have happened in the country in the past 3-5 years?
9. Are there data, policy briefs, national strategic plans or other sources available that quantify the changes made following the Optima HIV study?

**Dissemination of results**

10. Were results disseminated to key stakeholders?
    - What were the events (webinar, conference, workshop, stakeholder meetings, etc)?
    - Who were the key stakeholders?
    - How useful are these events?
    - Do you have any suggestions to improve the use of future study findings?
11. Was the presentation and policy brief provided clear enough to follow for policy makers?

**Feedback to improve future modelling exercise**

12. How satisfied was the NAP/NTP with the process and the support provided?
    - What would you have changed about the process and why?
        - Prompt: Study conceptualization, Involvement with in-country stake holders, the final products (PPT slide deck, brief/ report)
    - What types of questions did not get answered for you in the Optima application although you hoped to get answers?

13. Would you use the model for decision-support in future?

### Interview guide: Financing organizations

14. What feedback have you received from countries regarding the Optima analyses?
    - Prompts: perceived usefulness, received technical support, comprehension, choice of stakeholders
15. What effect (if any) did the Optima application have?
    - Supporting strategic thinking?
    - Supporting decision-making on financing?
    - Providing evidence for advocacy?
    - Identify and/or address data/evidence gaps?

16. Have funding applications been informed by Optima results?
    - Did the modelling projections guide other planning?
    - Did the study results improve policy makers' understanding of epidemic trends (historical trends; projected trends?
    - Any feedback on the accuracy of the projections/ estimates (trusting, perceptions)?
